# Subjective word-finding complaints in older adults: Development and preliminary psychometric evaluation of a novel self-reported questionnaire

**DOI:** 10.64898/2026.09.11.26362870

**Authors:** Fahed El-Khaldi, Anna Marier, Paolo Vitali, PREVENT-AD Research Group, Sylvia Villeneuve, Maxime Montembeault

## Abstract

**Objectives:** Subjective word-finding complaints (SWFC) are among the most frequent cognitive complaints in cognitively unimpaired older adults (CU). However, no comprehensive tools currently exist to assess this phenomenon. This study aimed to conduct a preliminary psychometric evaluation of a novel SWFC questionnaire, better characterize SWFC, and establish their demographic/clinical correlates.

**Methods:** 287 older adults (192 CU, 95 with mild cognitive impairment (MCI)) from the PREVENT-AD cohort completed a novel 29-item SWFC questionnaire available in French and English. Internal consistency and construct validity were evaluated. Relationships of demographic/clinical variables with total and domain-specific SWFC scores were analyzed.

**Results:** Internal consistency was strong (Cronbach’s Alpha=0.926). Difficulty remembering the names of famous people was the most frequently endorsed item (83%), whereas items related to the social impacts of SWFC were least frequently endorsed. Exploratory factor analysis revealed six distinct factors: SWFC coping strategies, fluency difficulties and affective impacts, SWFC-related errors, proper noun retrieval difficulties, social impacts of SWFC, and common noun retrieval difficulties. Older age was associated with higher total SWFC and greater proper and common noun retrieval difficulties, female sex with greater proper noun retrieval difficulties, higher education with lesser social impacts of SWFC, and MCI status with greater social impacts of SWFC.

**Discussion:** These findings demonstrate that SWFC are multidimensional rather than a unitary construct in aging. This novel questionnaire provides a promising tool for characterizing the type and severity of specific SWFC and may facilitate future research examining which complaint profiles are associated with adverse clinical outcomes, including Alzheimer’s disease.

## 1. Introduction

Alzheimer’s disease (AD) represents a major public health crisis. Typical AD dementia is characterized by cognitive changes in memory and at least one other domain (e.g., language, executive function, visuospatial abilities, or behavior), along with functional impairments^1^. Importantly, by the time clinical symptoms emerge, AD pathology, primarily amyloid accumulation, has often been developing in the brain for up to two decades^2^. This extended preclinical phase has led researchers and clinicians to focus on earlier stages of the disease, including mild cognitive impairment (MCI), in which subtle cognitive deficits are detectable but functional independence is largely maintained^1^.

Even earlier, subjective cognitive decline (SCD) has emerged as a promising concept for the older adult population, where many worry over a perceived decline in their cognitive abilities despite having normal scores on objective measures^3^. SCD has been associated with increased risk of progression to MCI^4,5^; elevated presence of AD biomarkers^6–8^; and brain atrophy^9–13^. While most studies assessing SCD have been centered around memory related complaints^14^, individuals with SCD may also report other subjective complaints independent of memory-related complaints, or alongside them.

For instance, subjective language complaints, and most precisely, subjective word-finding complaints (SWFC), have received more attention recently, as they rank as one of the most frequent and severe complaints among cognitively unimpaired older adults (CU), surpassing most memory complaints^5,15–18^. SWFC are characterized by difficulties such as forgetting the names of objects/people/places, finding the right words to use in a conversation or experiencing tip-of-the-tongue phenomenon^19^. Despite their frequency among CU, SWFC in the context of early AD detection is vastly underexplored as it has sometimes been considered as a normal part of ageing due to their prevalence. Nonetheless, it has been shown that SWFC were associated with lower concentrations of plasma Aβ1-42/Aβ1-40, gray matter atrophy in the fusiform gyrus and left Rolandic operculum^16^, as well as higher tau PET accumulation in inferior temporal regions^20^ in CU. Moreover, higher SWFC are associated with lower quality of life in AD dementia patients, more specifically in relationship to participation and enjoyment in social activities^21^.

Despite the high prevalence of SWFC and their emerging association with adverse clinical outcomes, current assessments are either often limited to single-item measures that lack psychometric robustness and granularity or focus on the consequences of word-finding difficulties rather than its frequency and how it manifests^16,22,23^. Conceptually, this may overlook the multidimensional nature of SWFC, which may manifest in diverse ways, for instance, difficulties retrieving common versus proper nouns, distinct error types such as hesitations or compensatory strategies, and varying emotional and social repercussions. Relying on such limited measures precludes the establishment of empirically derived thresholds at which SWFC become clinically significant. Given their high prevalence among older adults, SWFC are unlikely to represent a binary phenomenon. A comprehensive, multidimensional assessment is therefore necessary to identify when SWFC become clinically relevant and clarify current inconclusive evidence regarding demographic correlates^5,16,24,25^. While research investigating cognitive complaints and language complaints have found associations with correlates such as age, sex, and depression^18,24^, evidence specific to SWFC remains limited, and inconclusive.

Therefore, the aim of the present study is to develop and conduct a preliminary psychometric evaluation on a questionnaire designed to comprehensively capture SWFC in older adults. We expect the new scale to demonstrate good psychometric reliability, validity, and effectively capture the diverse manifestations of SWFC. In addition, we will examine associations between SWFC and demographic (age, sex, years of education, multilingualism (monolinguals vs multilinguals), and clinical (cognitive status (CU vs MCI)) characteristics. We anticipate that our comprehensive SWFC questionnaire will help in elucidating demographic and clinical correlates of SWFC, such as more elevated SWFC in older adults, in people with lower education and in MCI vs CU participants.

## 2. Methods

### 2.1 Participants

Participants were recruited from the Pre-symptomatic Evaluation of Experimental or Novel Treatments for Alzheimer’s Disease (PREVENT-AD) cohort, located in Montreal, Quebec, Canada. This is a longitudinal observational cohort, organized by the Stop-AD Centre (https://www.centre-stopad.com/en/), with the goal of detecting and tracking AD biomarkers in asymptomatic but at-risk older adults from the larger community.

Participants were initially enrolled into the cohort between 2011 and 2017. To be eligible for the cohort, participants had to: i) be aged 60 years or older and have at least one parent, or a minimum two siblings who have been diagnosed with AD; with exception to participants aged 55-59, who were eligible if their age was within 15 years of the symptom onset of their youngest affected relative ii) have at least 6 years of formal education; iii) be able to participate in scheduled regular visits iv) provide informed consent for multimodal assessments v) show an absence of cognitive impairment, which was confirmed using the Montreal Cognitive Assessment (MoCA ≥ 26/30), and the Clinical dementia rating (CDR = 0), vi) have a clinician confirm that they are cognitively normal through exhaustive neuropsychological assessment if their MoCA < 26, CDR > 0, or their Repeatable Battery for the Assessment of Neuropsychological Status (RBANS) score was below normal at baseline^26,27^.

It is important to note that although PREVENT-AD is a longitudinal cohort, the present study is cross-sectional in nature. The SWFC questionnaire was administered to all eligible participants at a single timepoint in February 2025. All demographic and diagnostic variables reflect participants’ status at the time of questionnaire completion. The time elapsed since each participant’s initial PREVENT-AD enrollment was not controlled for as it was not considered to bear on SWFC. Furthermore, some participants’ cognitive status has changed since their initial enrollment and have since developed MCI or dementia. Therefore, to be included in the current study, participants had to i) be classified as CU or as an older adult having MCI ii) not be diagnosed with dementia; ii) have completed our new SWFC questionnaire.

The online SWFC questionnaire was initially distributed to 359 participants, of which 292 completed the questionnaire. Two participants were excluded because they met diagnostic criteria for dementia, and three additional participants were excluded as statistical outliers (i.e., scores deviating by more than 2.5 standard deviations from the mean of total SWFC). The final sample therefore comprised 287 participants.

### 2.2 Procedure

#### 2.2.1. Questionnaire development

Our goal was to design a tool, in both English and French, that comprehensively assessed SWFC and captured changes relative to self-reported functioning from a decade earlier. The instrument was designed to evaluate three domains: (i) whether individuals experienced SWFC, (ii) the specific types of word-finding difficulties reported, and (iii) the impact of these difficulties on daily functioning and social interactions. A larger number of items than ultimately necessary were included to allow identification of the most informative items for a future, shorter version of the questionnaire.

##### Item generation

Questions were generated using our clinical judgement and reviewing the existing literature on language changes in older adults with and without dementia^28–32^. In total, an initial pool of 25 questions was drafted.

##### Feedback from language experts

The initial questionnaire was revised based on expert feedback from four speech-language pathologists and language researchers. Following these revisions, the questionnaire was refined to 29 items, enhancing its clarity, comprehensiveness, accessibility, and conceptual validity.

##### Instructions

Given that perceived cognitive decline is a key criterion for the diagnosis of MCI and AD dementia^1^, participants are asked to rate their current level of SWFC compared to 10 years ago. We opted for a 6-point Likert scale to provide greater granularity and enhance the tool’s sensitivity in detecting varying complaint severity. The scale ranges from 0-5 (0=never; 1=rarely, 2=sometimes, 3=frequently, 4=very frequently, 5=always). Each point is clearly well defined to reduce ambiguity and eliminate subjective interpretation by participants.

##### Translation

The questionnaire was translated from English to French (see supplementary table 1) using standard forward and back translation to ensure both sets of questionnaires are conceptually equivalent. Discrepancies were reviewed and revised to preserve semantic content throughout both sets of questionnaires.

#### 2.2.2 Validation

The questionnaire was electronically distributed via Qualtrics to participants in the PREVENT-AD cohort mid-January 2025. Reminder emails were sent in late January 2025 to participants who had not yet completed the questionnaire, and the final submissions were received in mid-February. Demographic (age, sex, education, multilingualism) and clinical (cognitive status (CU, MCI), Mini Mental State Examination (MMSE) scores, Repeatable Battery for the Assessment of Neuropsychological Status (RBANS) total scores, RBANS delayed memory recall scores, and RBANS language index scores) data collected within 12 months were then linked to each participant’s SWFC data.

### 2.3. Statistical Analyses

Total and item-level SWFC means were calculated across all 29 questionnaire items to determine the most frequently endorsed word-finding complaints.

#### 2.3.1 Psychometric evaluation of the questionnaire

Internal consistency of the overall questionnaire was evaluated using Cronbach’s alpha. Inter-item correlation and item-total correlation coefficients between items were calculated using Pearson’s r. If inter-item correlation coefficients exceeded 0.90, they were deemed redundant and would be excluded^33^. Data normality was examined using visual inspection and with a Shapiro-Wilk test. Exploratory factor analysis (EFA) was used to examine the latent structure of the questionnaire and identify subdomains that reflect distinct aspects of SWFC. For items that exhibited cross-loadings, each item was assigned to the factor on which it had the highest loading, with the exception of items whose content suggested a better conceptual fit with a different factor regardless of a higher cross-loading.

#### 2.3.2 Associations between demographic/clinical variables and SWFC score

To assess how demographic and clinical variables are associated with total SWFC scores, a set of analyses of covariance (ANCOVA) were conducted with total SWFC score as the dependent variable. The independent variables of interest are age (in years), sex (male vs female), education (in years), multilingualism (monolinguals vs multilinguals), cognitive status (CU vs MCI), MMSE scores, RBANS total score, RBANS delayed memory recall scores, and RBANS language index scores. For each ANCOVA model, one of these variables was entered as the independent variable, while age, sex, education, cognitive status, and test language (French vs English) served as covariates. Because the questionnaire was administered in both French and English, test language was included as a covariate in all models to account for potential differences between versions. As an additional check, sample statistics between the French and English subsamples were compared (supplementary table 2). For the binary independent variables (sex, cognitive status, and multilingualism), covariate-adjusted SWFC scores were obtained from the corresponding ANCOVA models and used to generate residualized mean box plots that compared both groups.

To extend these analyses to the latent components that were derived using EFA, a multivariate analysis of covariance (MANCOVA) was conducted with the sum of scores of each EFA-derived factor scores serving as the dependent variable. Sum of scores were converted to z-scores as each factor had a different number of items that resulted in different total scores. Age, sex, education, cognitive status, and multilingualism were entered as independent variables, while age, sex, education, cognitive status, and test language served as covariates. Follow up univariate analysis of covariance (ANCOVA) were computed to identify specific sources of multivariate effects.

Partial correlations were calculated primarily to produce plots shown in the figures. These correlations were obtained using ordinary least squares (OLS) regression within a two-step residualization procedure that removed the effects of age, sex, education, and cognitive status. The general model was:

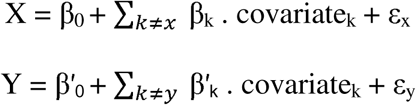

Residuals from these models were used to calculate the partial correlations relating age, sex, education, and cognitive status to either total SWFC score or PCA-derived factor scores.

## 3. Results

### 3.1. Descriptives

287 participants were included in the study (198 females, 89 males; see table 1). Of these, 192 were classified as CU, and 95 as having MCI. The mean age of all participants was 71.50 (SD =5.64). Most participants were highly educated (M=15.85, SD = 3.38) and global cognition as assessed with the MMSE (M=28.73, SD =1.43) and the RBANS total score (M=99.93, SD = 10.17) were within normal range. The mean SWFC total score for the whole sample was 24.89 (SD = 12.46).

**Table 1:**
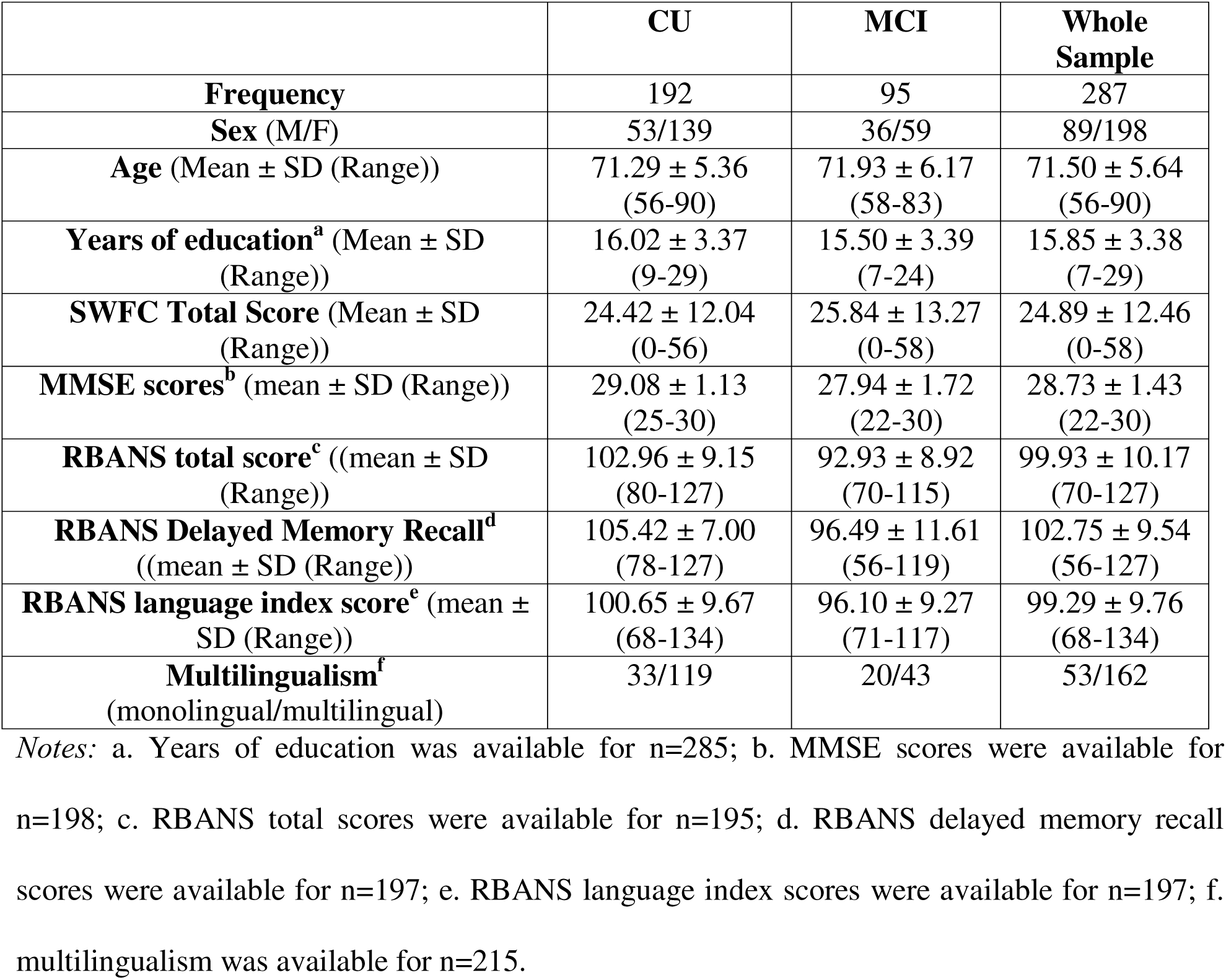
Sample demographics.

|  | **CU** | **MCI** | **Whole Sample** |
| --- | --- | --- | --- |
| **Frequency** | 192 | 95 | 287 |
| **Sex (M/F)** | 53/139 | 36/59 | 89/198 |
| **Age (Mean ± SD (Range))** | 71.29 ± 5.36<br>(56-90) | 71.93 ± 6.17<br>(58-83) | 71.50 ± 5.64<br>(56-90) |
| **Years of education<sup>a</sup> (Mean ± SD (Range))** | 16.02 ± 3.37<br>(9-29) | 15.50 ± 3.39<br>(7-24) | 15.85 ± 3.38<br>(7-29) |
| **SWFC Total Score (Mean ± SD (Range))** | 24.42 ± 12.04<br>(0-56) | 25.84 ± 13.27<br>(0-58) | 24.89 ± 12.46<br>(0-58) |
| **MMSE scores<sup>b</sup> (mean ± SD (Range))** | 29.08 ± 1.13<br>(25-30) | 27.94 ± 1.72<br>(22-30) | 28.73 ± 1.43<br>(22-30) |
| **RBANS total score<sup>c</sup> ((mean ± SD (Range))** | 102.96 ± 9.15<br>(80-127) | 92.93 ± 8.92<br>(70-115) | 99.93 ± 10.17<br>(70-127) |
| **RBANS Delayed Memory Recall<sup>d</sup> ((mean ± SD (Range))** | 105.42 ± 7.00<br>(78-127) | 96.49 ± 11.61<br>(56-119) | 102.75 ± 9.54<br>(56-127) |
| **RBANS language index score<sup>e</sup> (mean ± SD (Range))** | 100.65 ± 9.67<br>(68-134) | 96.10 ± 9.27<br>(71-117) | 99.29 ± 9.76<br>(68-134) |
| **Multilingualism<sup>f</sup> (monolingual/multilingual)** | 33/119 | 20/43 | 53/162 |

Notes: a. Years of education was available for n=285; b. MMSE scores were available for n=198; c. RBANS total scores were available for n=195; d. RBANS delayed memory recall scores were available for n=197; e. RBANS language index scores were available for n=197; f. multilingualism was available for n=215.

Figure 1 presents the mean severity ratings for each questionnaire item. Twelve items had a mean score of ≥1.0, indicating they were at least minimally endorsed (i.e., experienced "rarely" or more frequently) by older adults. Notably, the most highly endorsed item involved proper noun retrieval (e.g., difficulty remembering the names of famous people), in which 83% of participants reported a score of 2 (‘sometimes’) or higher on the scale. Items pertaining to the tip-of-the-tongue phenomenon (76%), forgetting the names of people they know but are not close with (68%), finding the right word to use in a conversation (67%) forgetting the names of objects (48%), being frustrated by SWFC (43%), pausing (41%), requesting help with SWFC (39%), and hesitating (35%) were also highly endorsed. In contrast, most items related to the psychological or social impact of SWFC (e.g., avoiding social activities (2%), being infantilized (<1%)) were less frequently endorsed. A full breakdown of item-response distribution is available in supplementary Figure 1.

**Figure 1:**
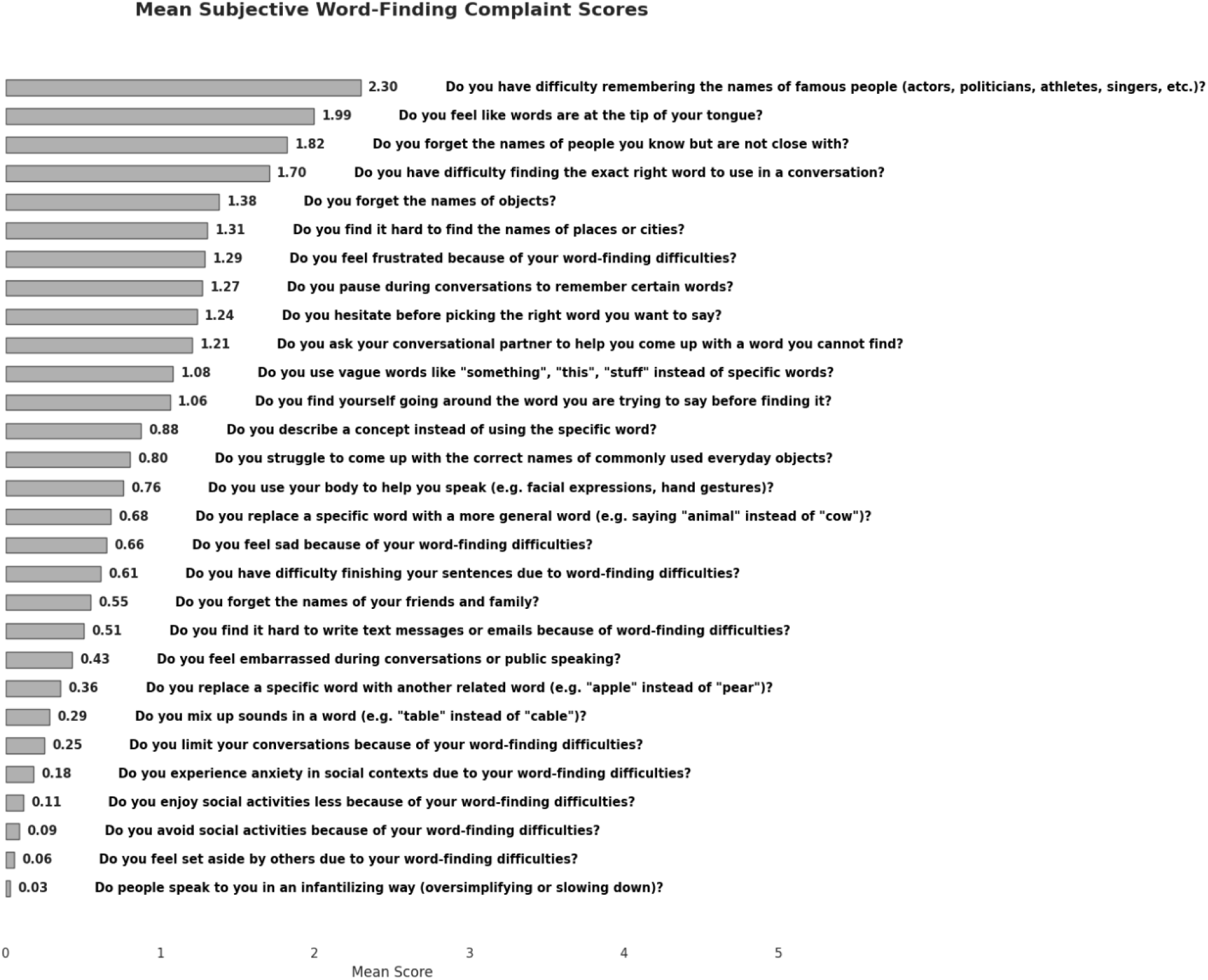
Mean scores for each item on the SWFC questionnaire (n=287). Items are ordered from the highest to the lowest mean, reflecting which word-finding difficulties are most and least endorsed by older adults. Scores range from 0-5 (0=never; 1=rarely, 2=sometimes, 3=frequently, 4=very frequently, 5=always), with higher values indicating greater frequency and severity of the complaint. Alt text: Mean score of each SWFC item sorted in descending order, with higher scores meaning that participants reported that they had these complaints more often.

### 3.2. Psychometric properties of the questionnaire

The scale demonstrated good internal consistency, with a Cronbach’s alpha of 0.926, supporting its reliability and cohesion. Supplementary figure 2 shows inter-item correlations ranging from .02 to .74, with most falling in the moderate range (0.25 - 0.50). Some pairs showed weak associations (e.g., items 6 and 28, r = 0.02), while others showed strong inter-item correlations (e.g. items 25 and 26, r = 0.74). No inter-item correlations exceeded 0.90, and thus all the items were retained. All items correlated at least moderately with the total score (r = 0.24 - 0.72).

Supplementary Figure 3 shows the normality distribution of the questionnaire. Based on visual inspection, the questionnaire shows a normal distribution. However, based on statistical analysis, the questionnaire slightly deviated from a normal distribution (Shapiro-Wilk W=0.972, p > 0.001) with a slightly platykurtic distribution (kurtosis= −0.244) that slightly skews to the right (skewness= 0.514).

EFA using the Kaiser criterion (eigenvalues > 1) suggested a six-factor solution. After examining the scree plot and considering clinical interpretability, we loaded the items onto six factors (refer to table 2). The six-factor model accounted for 61.1% of the total variance. The factors yielded clinically meaningful factors. The first factor included items related to SWFC coping strategies, which are the behavioural responses people use when experiencing SWFC (items 6,12,13,19,20). The second factor included items related to fluency difficulties and affective impacts (items 8,10,14,21,22,23,34,29). The third factor included items relating to SWFC-related errors (items 9,15,16,17,18). The fourth factor included items related to proper noun retrieval difficulties (items 4,5,7,11). The fifth factor included items related to the social impacts of SWFC (25,26,27,28). The sixth factor included items related to common noun retrieval difficulties (items 1,2,3).

**Table 2:**
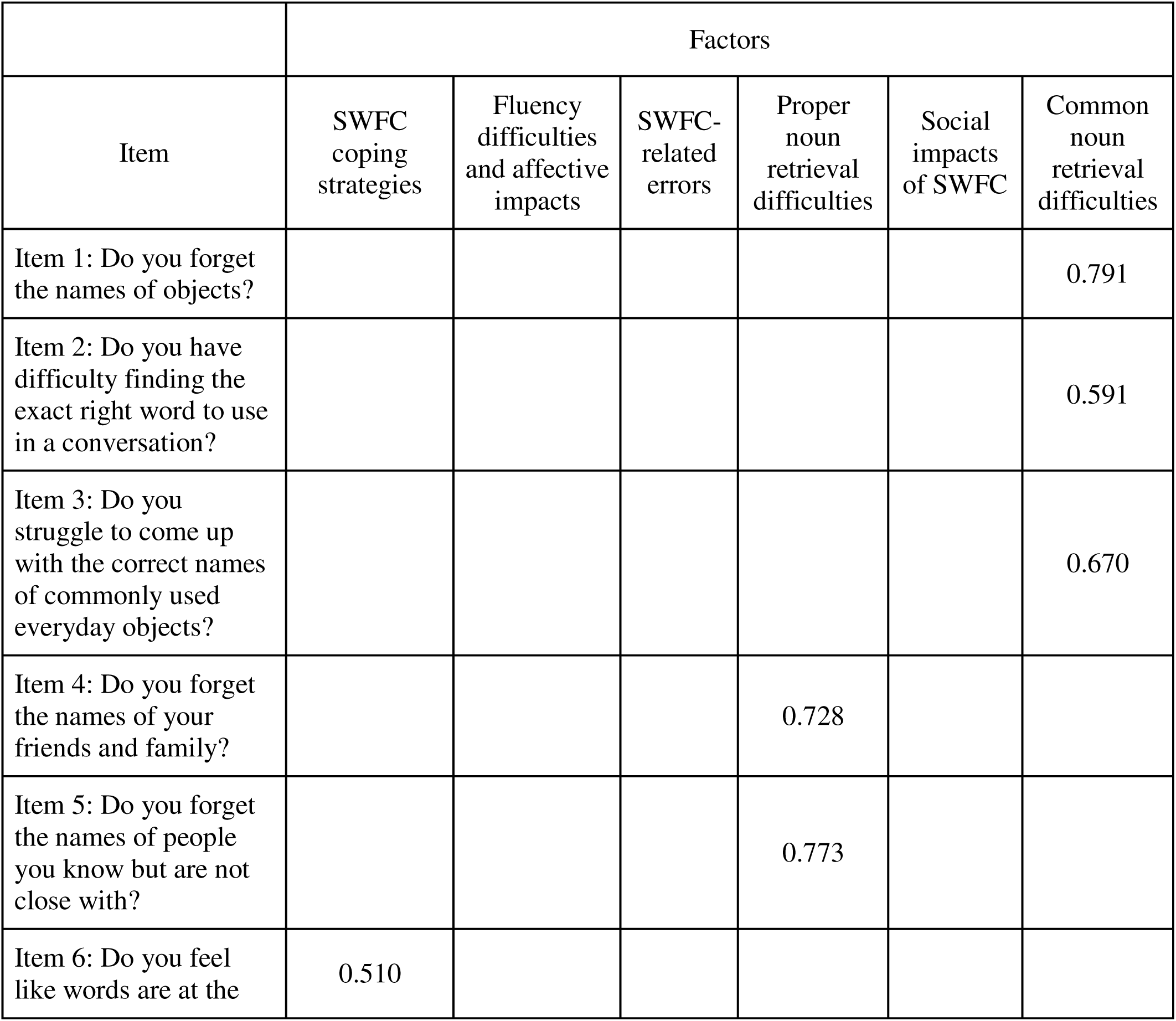
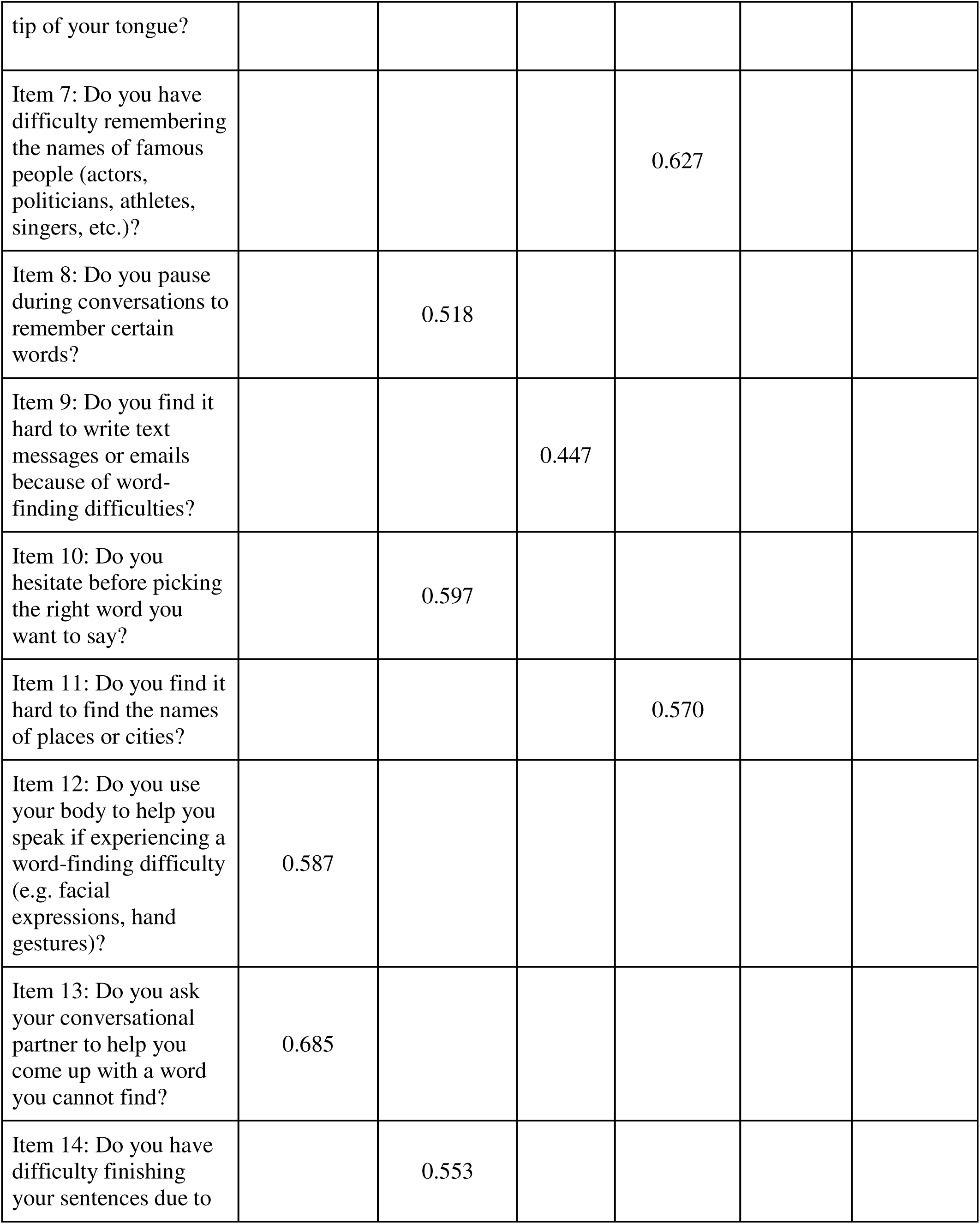
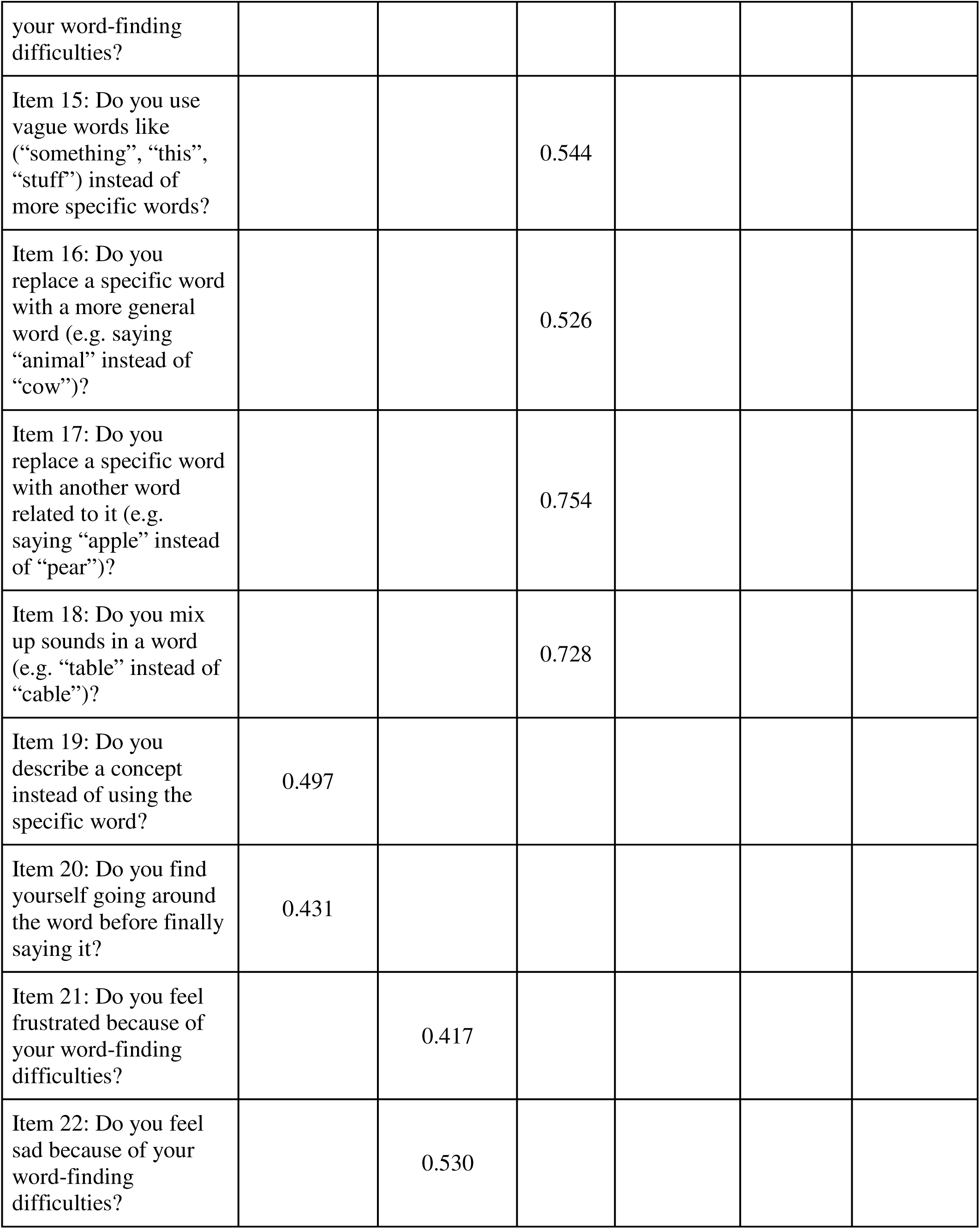
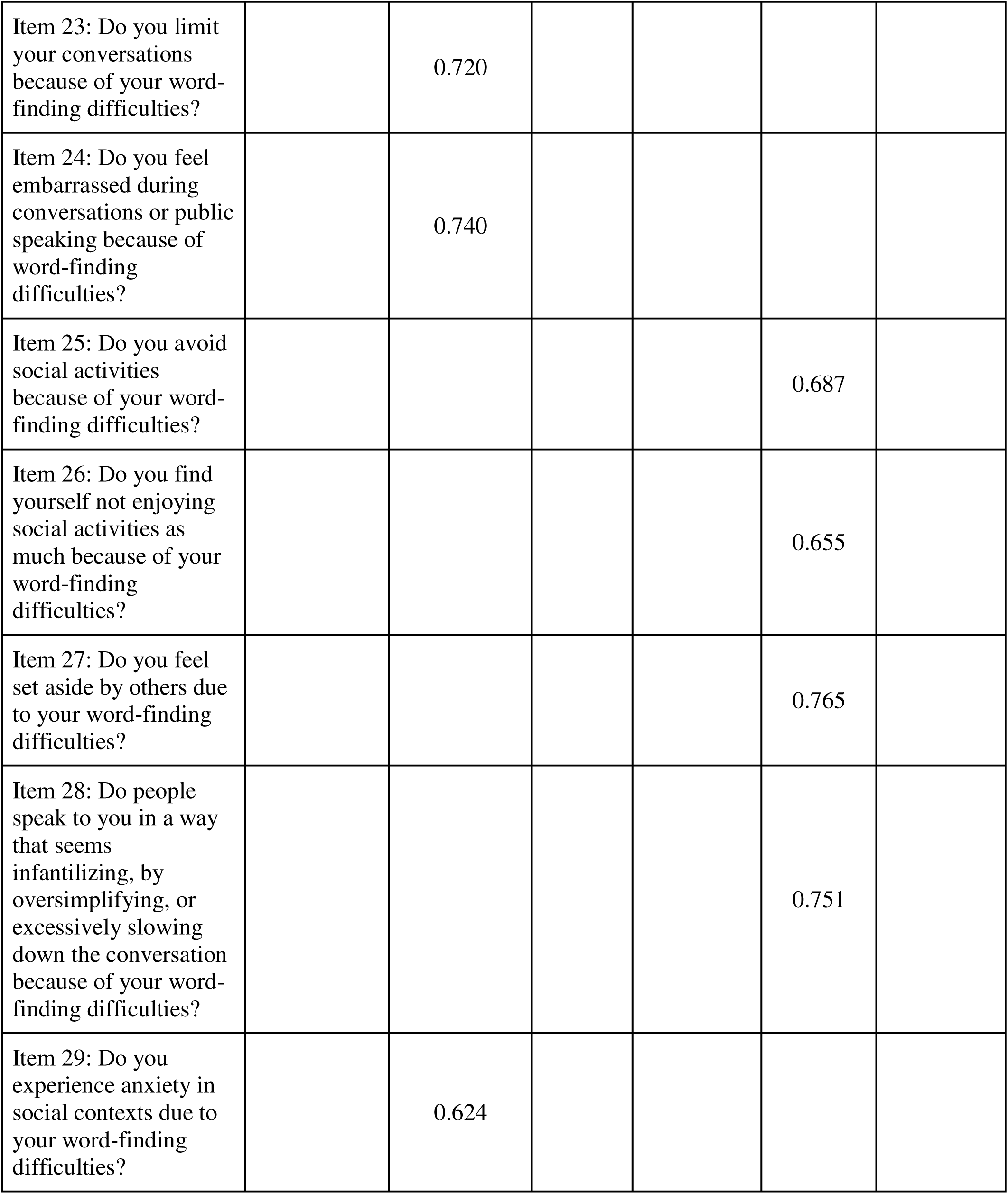
Factors corresponding to the scale’s items. Using Varimax rotation, six factors were extracted: Factor 1 contains items related to SWFC coping strategies; factor 2 contains items related to fluency difficulties and affective impacts; factor 3 contains items related to SWFC related errors; factor 4 contains items related to proper noun retrieval difficulties; factor 5 contains items related to the social impacts of SWFC; factor 6 contains items related to common noun retrieval difficulties.

### 3.3. Associations between demographics/clinical variables and SWFC scores

Figure 2 shows the residualized regression plots after adjusting for covariates (age, sex, education, cognitive status, and test language) for the associations between total SWFC scores and demographic/clinical variables. There was a significant positive correlation between age and SWFC total scores (r = 0.15, p = 0.010), where older participants were more likely to have higher SWFC total scores. No significant associations were found for education, MMSE scores, RBANS total scores, RBANS delayed memory recall scores, RBANS language index scores, multilingualism (monolingual vs bilingual), sex (males vs females), and cognitive status (CU vs MCI).

**Figure 2:**
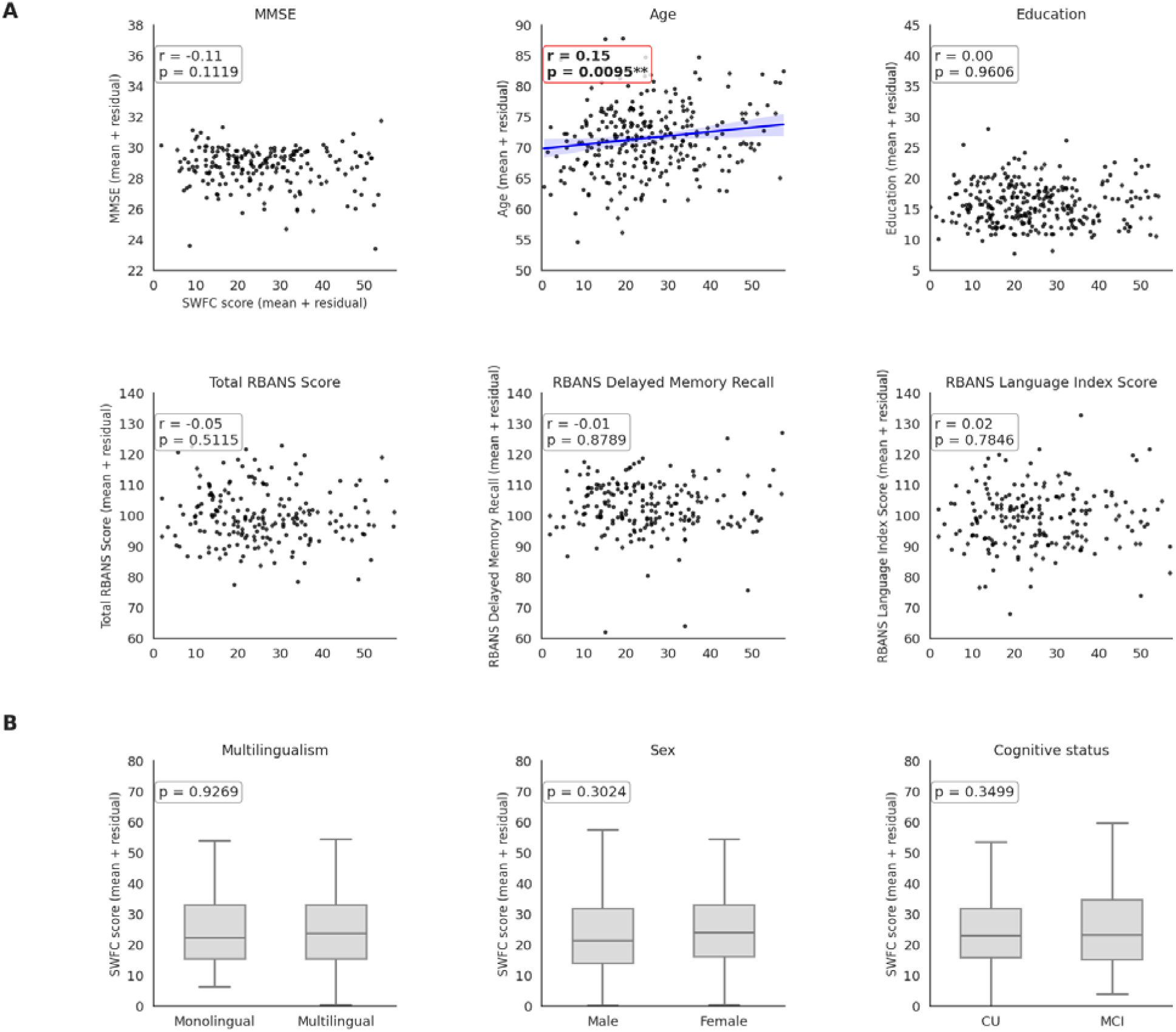
Associations of cognitive status and demographic variables with total SWFC score. Variables were regressed using Ordinary least squares (OLS) regression to obtain residuals that represent variance unexplained by covariates (age, sex, education, cognitive status, test language). Using these residuals for each variable, the partial correlations were then plotted. Significant positive associations were found between age and total SWFC score where the older a participant is, the higher their total SWFC score. Residualized mean scores for education, MMSE scores, RBANS total scores, RBANS delayed memory recall scores, RBANS language index scores, multilingualism, cognitive status and sex revealed no significant effects. Asterisks denote statistical significance (*p < .05, **p < .01, **p < .001). Alt text: Scatterplots showing correlations between total SWFC scores and each demographic (age, sex, education, multilingualism) or cognitive (MMSE score, cognitive status, RBANS-related measures) variable after adjusting for the other demographic and cognitive variables.

A MANCOVA examined the multivariate effects of the demographic/clinical variables on the six SWFC factors. There were significant multivariate effects of education (Wilks’ Λ = 0.937, F(6, 274) = 3.08, p = 0.006, partial η^2^ = 0.063), and sex (Wilks’ Λ = 0.936, F(6, 274) = 3.10, p = 0.006, partial η^2^ = .064). No significant multivariate effects were found for age, cognitive status, and multilingualism.

Figure 3 shows the residualized regression plots after adjusting for covariates (age, sex, education, cognitive status, and test language) for the associations between each SWFC factor and demographic/clinical variables, with the full statistics from the follow-up univariate ANCOVAs reported in supplementary table 3. Older participants were more likely to report more proper noun retrieval difficulties (F(1, 279) = 6.90, p = 0.010, partial η^2^ = 0.024), and general SWFC (F(1, 279) = 7.80, p = 0.006). More educated participants were more likely to report fewer social impacts of SWFC (F(1, 279) = 4.21, p = 0.041, partial η^2^ = 0.015). Females were more likely to report higher proper noun retrieval difficulties (F(1, 279) = 7.28, p = 0.007, partial η^2^ = 0.025). MCI participants were more likely to report more social impacts of SWFC (F(1, 279) = 6.28, p = 0.013, partial η^2^ = 0.022). No significant predictors emerged for SWFC coping strategies, fluency difficulties and affective impacts, or SWFC-related errors.

**Figure 3:**
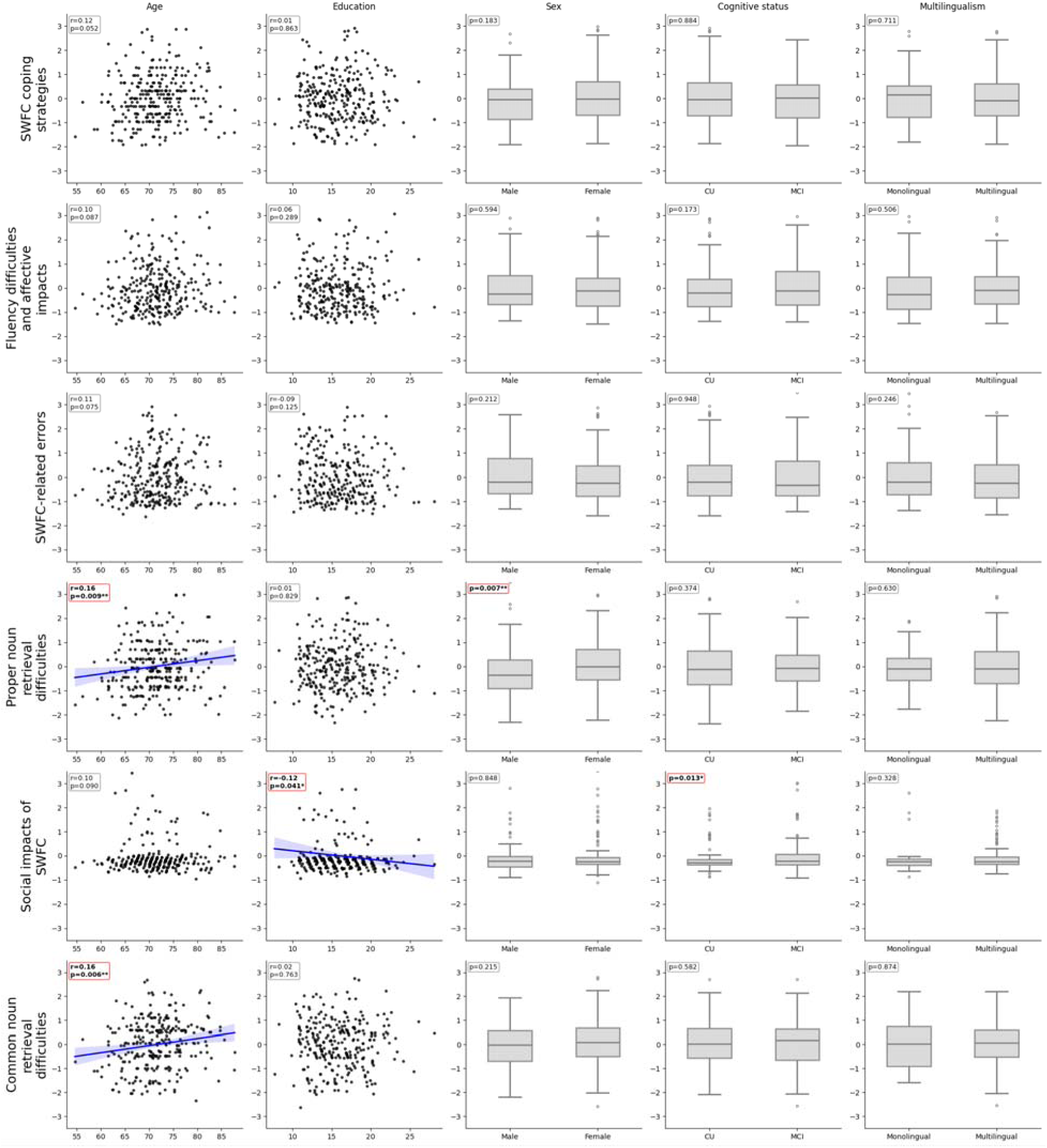
Associations of demographic/cognitive variables with SWFC factor scores. Relationships between demographics and cognitive status (represented in each column as age, sex, education, cognitive status, and multilingualism) and the six EFA-derived factors of SWFC (represented across each row, with factor 1 on SWFC coping strategies; factor 2 on fluency difficulties and affective impacts; factor 3 on SWFC related errors; factor 4 on proper noun retrieval difficulties; factor 5 on the social impacts of SWFC; factor 6 on common noun retrieval difficulties) were assessed after adjusting for covariates (age, sex, education, cognitive status, and test language). Continuous variables were plotted as partial correlations, and box plots represent the adjusted group mean differences for the categorical variables (sex and cognitive status: CU= cognitively unimpaired; MCI = mild cognitive impairment). Values represent partial correlations (*p <0.05; **p < 0.01), and the standard mean errors are represented in error bars. Alt text: Multi panel scatterplots depicting associations between the six derived factors and each demographic and cognitive variable after adjusting for age, sex, education, cognitive status, and test language.

## 4. Discussion

The aim of this study was to develop and conduct a preliminary psychometric evaluation on a questionnaire to comprehensively capture SWFC in older adults. Unlike previous measures, which relied on single-item measures or questionnaires limited to the functional impacts of anomia in clinical populations, our questionnaire offers a comprehensive assessment of SWFC experienced by older adults in the general population. It confirmed that SWFC are elevated and widespread among older adults, demonstrates good reliability and validity, and yields a well-distributed range of scores in the CU/MCI population. By capturing the multidimensional nature of SWFC with greater sensitivity, it revealed six distinct and clinically meaningful domains. Finally, it is correlated with relevant demographic and clinical variables: Common noun and proper noun retrieval difficulties increased with age; proper noun retrieval difficulties were more pronounced in females; higher education was associated with fewer social impacts of SWFC; and MCI participants reported greater social impacts of SWFC than CU participants. The novel SWFC questionnaire will facilitate research aimed at identifying thresholds associated with adverse outcomes, including biomarker positivity and clinical progression, and will help clarify the role of SWFC in the context of preclinical Alzheimer’s disease.

The questionnaire allowed us to characterize how SWFC manifest in older adults. Specifically, the factor structure indicates that SWFC encompass six dimensions: (1) SWFC coping strategies (2) fluency difficulties and affective impacts, (3) SWFC-related errors, (4) proper noun retrieval difficulties, 5) social impacts of SWFC, and 6) common noun retrieval difficulties. Items involving proper nouns emerged as some of the most frequently endorsed SWFC among CU (>67% endorsed these items), consistent with previous studies identifying proper name retrieval as a common complaint in aging^15,34^. This pattern aligns with a substantial literature on age-related proper name retrieval deficits, which has shown that older adults experience more frequent retrieval failures specifically for proper names compared to common nouns^19,35^. A likely explanation is rooted in the structure of semantic networks. Common nouns are embedded in rich networks with multiple semantic neighbors and overlapping features, which can support compensatory retrieval strategies and tolerated substitutions. For example, a sofa or a couch may be approximated as a “chair” without loss of communicative intent. In contrast, proper nouns refer to unique entities and have minimal semantic connectivity, offering no acceptable substitute when retrieval fails^36^. Because proper names have weaker semantic and associative pathways, they are particularly vulnerable to aging effects on lexical access^37^. An alternative explanation is that proper nouns may be more salient or emotionally meaningful, making retrieval failures more noticeable and therefore more likely to be reported, independent of objective frequency. Future work combining subjective reports with naming tasks could help clarify these mechanisms. In contrast, items assessing social impacts were the least endorsed in our sample. Nonetheless, frustration related to SWFC was frequently reported, suggesting that older adults may currently compensate for their difficulties while remaining aware of and bothered by them. Frustration may therefore represent a subtle change in language functioning before it translates into measurable functional impairment, consistent with models positing affective responses as a possible risk factor of future cognitive decline^38^.

The second aim of this study was to examine associations between SWFC and demographic or clinical characteristics. Previous research based on a single SWFC questionnaire item reported no associations with sex, age, education, depression or anxiety-related symptoms^16^. By capturing the multidimensional nature of SWFC, the questionnaire proved more sensitive to individual variability than previous single-item measures. Our analysis uncovered distinct demographic profiles: Older adults were more likely to report proper noun retrieval difficulties, a pattern that may be consistent with the generally higher age-related risk of AD^39^. Sex was not associated with total SWFC scores, but females reported more proper noun retrieval difficulties at the factor level. While the literature on sex differences in subjective cognitive complaints is mixed and inconclusive^24,25,40–42^, females tend to report more worries associated with their SCD^43^. Education showed a comparable pattern: total SWFC scores did not vary with years of education, but individuals with higher education reported fewer social impacts of SWFC. This may reflect increased cognitive self-monitoring or hypernosognosia in highly educated people CU^44^, or heightened attention to more socially salient complaints such as proper noun retrieval difficulties among females. CU and MCI participants reported similar overall SWFC, but MCI participants endorsed greater social impacts of SWFC. This is consistent with early functional consequences along the AD spectrum and provides preliminary support for the clinical relevance of the scale. In line with the clinical description of MCI, this finding suggests that, at this stage, SWFC may begin to interfere with communication and social interactions. At the MCI stage, objective word-finding changes are detectable, such that reported SWFC may reflect genuine word-finding failures rather than isolated subjective experiences^45^. These word-finding failures may, in turn, contribute to the greater social impacts of SWFC reported by individuals with MCI. By contrast, SWFC were not associated with MMSE and RBANS-related measure scores, likely reflecting ceiling effects and conceptual differences between global cognitive screening and subjective language complaints. Together, these findings help refine the profile of individuals more likely to experience SWFC and highlight the value of using a comprehensive tool to capture subtle, subjective word-finding difficulties.

The development and validation of this comprehensive SWFC questionnaire have several important research and clinical implications. In the short term, the tool will enable more precise characterization of SWFC in aging by capturing multiple dimensions of the phenomenon rather than relying on single-item measures. This work builds directly on recent evidence showing that SWFC are associated with AD pathology: Given that SWFC correlated with CSF amyloid and tau PET accumulation in CU^16,20^, SWFC may reflect early AD-related processes. The present scale provides the opportunity to examine these relationships more rigorously by going beyond a single item and assessing whether specific SWFC domains map onto AD biomarkers such as CSF and PET measures, as well as emerging scalable plasma markers such as p-tau217. These findings could also motivate longitudinal studies assessing whether SWFC predict conversion to MCI in the context of AD pathology and help determine whether specific facets of SWFC signal early functional vulnerability. In the longer term, item-level analyses could inform the development of a shorter, clinically oriented version of the questionnaire containing only the most predictive items, providing a low-cost, non-invasive screening tool for settings where biomarker testing is limited.

While these results provide valuable insight into SWFC in older adults, they should be interpreted in light of several limitations. First, personality traits, mood, and depressive symptoms were not assessed, even though higher neuroticism and depressive symptoms are associated with greater subjective cognitive complaints^46^, suggesting that affective factors may influence SWFC reporting. Future studies should clarify whether SWFC reflects emotional mechanisms in addition to semantic difficulty. Second, the cohort covered a relatively restricted age range (56–90), which may limit detection of early age-related changes; including younger older adults will help determine when SWFC first emerge. Third, PREVENT-AD primarily consists of highly educated, white older adults, limiting generalizability and potentially reducing sensitivity to demographic effects. Fourth, MCI participants were very mild (mean MMSE = 27.9), which may have reduced our ability to detect differences between CU and MCI; replication in more diverse cohorts with broader cognitive variability will help determine whether SWFC more clearly distinguishes early clinical stages of the AD spectrum. Fifth, because the questionnaire was administered in both French and English, formal measurement invariance testing across the two language versions was not feasible given the small English-version subsample; establishing invariance in a larger sample remains an important direction for future work. Finally, confrontation naming performance was not included in our analyses as it was available only as a raw, unstandardized score; examining the association between SWFC and confrontation naming using an appropriately normed measure (such as the Boston Naming Test) is another important direction for future work. Taken together, these results should be interpreted as preliminary evidence derived from a single selected cohort. Since our sample consisted of predominately of highly educated white older adults, this limits the generalizability of our findings to broader, more diverse clinical and community populations where people may differ in educational background, race and ethnicity, and clinical severity. Because external validity is a key foundation for any newly developed tool, establishing the questionnaire’s psychometric properties and applicability across more cohorts is an essential next step before it can be recommended for wider clinical or community use.

These findings highlight the importance of examining language, and particularly SWFC alongside memory in the study of cognitive aging. Word-finding concerns are among the most frequent cognitive complaints in older adults, yet they receive comparatively less attention in research and clinical practice. By developing a multidimensional questionnaire, this study provides a clearer way to capture the specific forms these complaints take and moves beyond single-item assessments. Given their prevalence, reliable tools to measure SWFC are essential to determine whether they hold clinical meaning, including links to biological markers and future cognitive outcomes. If certain aspects of these complaints predict adverse trajectories, they may offer a scalable and personally relevant signal to guide next steps for older adults, including earlier counselling, monitoring, and support. More broadly, recognizing language changes as a central feature of cognitive aging supports a more complete and person-centered understanding of decline, ensuring that concerns most salient to older adults are appropriately valued in research and care.

## Data Availability

All data produced in the present work are contained in the manuscript

## Acknowledgements

The authors acknowledge the PREVENT-AD staff and research group members for coordinating the distribution of the questionnaire and compiling the data used in this study. We thank the participants of PREVENT-AD for their time and effort in helping us collect this data. We also thank the four speech-language experts who provided valuable input and contributed to the creation of the questionnaire.

## Statements and Declarations

### Ethical Considerations

The PREVENT-AD study was conducted following the ethical standards of all involved institutional research committees and adheres to the principles outlined in the 1975 Helsinki Declaration and its later amendments. The study was approved by the CIUSSS-COMTL Research Ethics Board.

### Consent to Participate

Written informed consent to participate was obtained from all participants.

### Consent for Publication

Written informed consent for publication was obtained from all participants.

### Data Availability

The data on participants who agreed to sharing can be accessed via the PREVENT-AD Data Sharing Repository https://registeredpreventad.loris.ca/. Given that sensitive information is shared, access is restricted to qualified researchers or physicians. The PREVENT-AD coordinator will verify information of all principal investigators requesting an account before granting access, after which data can be shared with trainees who are under their supervision and for whom they take full responsibility for data usage. All terms of use must be agreed to when requesting an account, at https://registeredpreventad.loris.ca/ login/request-account/. In brief, users must not attempt to sell or claim intellectual property rights in the PREVENT-AD dataset, users must obtain any needed ethics approvals for their use of the PREVENT-AD dataset, the data must be used for “neuroscience research”, the data must not be redistributed, and any attempt to re-identify participants is forbidden. When used in publications, the methods section must state PREVENT AD as the source of the data and the manuscript describing the first open release4 or the current release must be cited depending on the data used.

## References

1. McKhann GM, Knopman DS, Chertkow H, et al. The diagnosis of dementia due to Alzheimer’s disease: recommendations from the National Institute on Aging Alzheimer’s Association workgroups on diagnostic guidelines for Alzheimer’s disease. Alzheimer’s & dementia. 2011;7(3):263–269.

2. Dubois B, Hampel H, Feldman HH, et al. Preclinical Alzheimer’s disease: Definition, natural history, and diagnostic criteria. Alzheimer’s & Dementia. 2016;12(3):292–323. 10.1016/j.jalz.2016.02.002

3. Jessen F, Amariglio RE, Van Boxtel M, et al. A conceptual framework for research on subjective cognitive decline in preclinical Alzheimer’s disease. Alzheimer’s & dementia. 2014;10(6):844–852.

4. Fernández-Blázquez MA, Ávila-Villanueva M, Maestú F, Medina M. Specific Features of Subjective Cognitive Decline Predict Faster Conversion to Mild Cognitive Impairment. Journal of Alzheimer’s Disease. 2016;52(1):271–281. doi:10.3233/jad-150956

5. Mazzeo S, Padiglioni S, Bagnoli S, et al. Assessing the effectiveness of subjective cognitive decline plus criteria in predicting the progression to Alzheimer’s disease: an 11 year follow up study. European Journal of Neurology. 2020;27(5):894–899.

6. Janssen O, Jansen WJ, Vos SJ, et al. Predictors of preclinical Alzheimer’s disease in persons with subjective cognitive decline: Neuropsychiatry and behavioral neurology/presymptomatic disease/prodromal disease/prodromal states. Alzheimer’s & Dementia. 2020;16:e042658.

7. Mengel D, Soter E, Ott JM, et al. Blood biomarkers confirm subjective cognitive decline (SCD) as a distinct molecular and clinical stage within the NIA-AA framework of Alzheimeŕ s disease. Molecular Psychiatry. 2025:1–10.

8. Perrotin A, La Joie R, de La Sayette V, et al. Subjective cognitive decline in cognitively normal elders from the community or from a memory clinic: differential affective and imaging correlates. Alzheimer’s & Dementia. 2017;13(5):550–560.

9. Chen B, Wang Q, Zhong X, et al. Structural and functional abnormalities of olfactory-related regions in subjective cognitive decline, mild cognitive impairment, and Alzheimer’s disease. International Journal of Neuropsychopharmacology. 2022;25(5):361–374.

10. Hu X, Teunissen CE, Spottke A, et al. Smaller medial temporal lobe volumes in individuals with subjective cognitive decline and biomarker evidence of Alzheimer’s disease—Data from three memory clinic studies. Alzheimer’s & Dementia. 2019;15(2):185–193.

11. Kuhn E, Perrotin A, La Joie R, et al. Association of the informant-reported memory decline with cognitive and brain deterioration through the Alzheimer clinical continuum. Neurology. 2023;100(24):e2454–e2465.

12. Kuhn E, Perrotin A, Tomadesso C, et al. Subjective cognitive decline: opposite links to neurodegeneration across the Alzheimer’s continuum. Brain Communications. 2021;3(3):fcab199.

13. Wang X, Wang M, Wang X, et al. Subjective cognitive decline-related worries modulate the relationship between global amyloid load and gray matter volume in preclinical Alzheimer’s disease. Brain Imaging and Behavior. 2022;16(3):1088–1097.

14. Bruus AE, Waldemar G, Vogel A. Subjective complaints are similar in subjective cognitive decline and early-stage Alzheimer’s disease when assessed in a memory clinic setting. Journal of Geriatric Psychiatry and Neurology. 2023;36(6):479–486.

15. Condret-Santi V, Barbeau E, Matharan F, Le Goff M, Dartigues J-F, Amieva H. Prevalence of word retrieval complaint and prediction of dementia in a population-based study of elderly subjects. Dementia and Geriatric Cognitive Disorders. 2013;35(5-6):313–324.

16. Montembeault M, Stijelja S, Brambati SM, Initiative FtAsDN. Self-reported word-finding complaints are associated with cerebrospinal fluid amyloid beta and atrophy in cognitively normal older adults. Alzheimer’s & Dementia: Diagnosis, Assessment & Disease Monitoring. 2022;14(1):e12274. 10.1002/dad2.12274

17. Diaz-Galvan P, Ferreira D, Cedres N, et al. Comparing different approaches for operationalizing subjective cognitive decline: impact on syndromic and biomarker profiles. Scientific Reports. 2021;11(1):4356.

18. Martins I, Mares I, Stilwell P. How subjective are subjective language complaints. European journal of neurology. 2012;19(5):666–671.

19. Burke DM, MacKay DG, Worthley JS, Wade E. On the tip of the tongue: What causes word finding failures in young and older adults? Journal of memory and language. 1991;30(5):542–579.

20. Marier A, Fernández Arias J, Aumont É, et al. Language deficits across PET based Braak stages of tau accumulation in Alzheimer’s disease. Alzheimer’s & Dementia. 2026;22(3):e71286.

21. Farrell MT, Zahodne LB, Stern Y, Dorrejo J, Yeung P, Cosentino S. Subjective word finding difficulty reduces engagement in social leisure activities in Alzheimer’s disease. Journal of the American Geriatrics Society. 2014;62(6):1056–1063.

22. Javaudin J. Réalisation d’une échelle de plainte concernant l’anomie chez les patients atteints de troubles neurologiques. 2021;

23. Joly F, Weisse S. Échelle de plainte concernant l’anomie: élaboration d’une version courte. 2022;

24. Pinho PJ, Bertola L, Ramos AA, et al. Subjective memory complaints: Prevalence, associated factors and sex differences in the ELSI Brazil study. International Journal of Geriatric Psychiatry. 2023;38(11):e6026.

25. Schliep KC, Barbeau WA, Lynch KE, et al. Overall and sex-specific risk factors for subjective cognitive decline: findings from the 2015–2018 Behavioral Risk Factor Surveillance System Survey. Biology of sex Differences. 2022;13(1):16.

26. Tremblay-Mercier J, Madjar C, Das S, et al. Open science datasets from PREVENT-AD, a longitudinal cohort of pre-symptomatic Alzheimer’s disease. NeuroImage: Clinical. 2021;31:102733.

27. Villeneuve S, Poirier J, Breitner JC, et al. The PREVENT AD cohort: Accelerating Alzheimer’s disease research and treatment in Canada and beyond. Alzheimer’s & Dementia. 2025;21(10):e70653.

28. Cavanaugh R, Haley KL. Subjective communication difficulties in very mild aphasia. American Journal of Speech-Language Pathology. 2020;29(1S):437–448.

29. Farias ST, Weakley A, Harvey D, Chandler J, Huss O, Mungas D. The measurement of everyday cognition (ECog): revisions and updates. Alzheimer Disease & Associated Disorders. 2021;35(3):258–264.

30. Macoir J, Laforce R, Wilson M, Tremblay M, Hudon C. The role of semantic memory in the recognition of emotional valence conveyed by written words. Aging, Neuropsychology, and Cognition. 2020;27(2):270–288.

31. Nutter-Upham KE, Saykin AJ, Rabin LA, et al. Verbal fluency performance in amnestic MCI and older adults with cognitive complaints. Arch Clin Neuropsychol. May 2008;23(3):229–41. doi:10.1016/j.acn.2008.01.005

32. O’Connor K. Stories of people with dementia who experience word-finding difficulties. University of East London; 2024.

33. Field A. Discovering statistics using IBM SPSS statistics 5th ed. Sage; 2018.

34. Schweich M, van der Linden M, Bredart S, Bruyer R, Nelles B, Schils JP. Daily-life difficulties in person recognition reported by young and elderly subjects. Applied Cognitive Psychology. 1992;6(2):161–172. doi:10.1002/acp.2350060206

35. Evrard M. Ageing and lexical access to common and proper names in picture naming. Brain and language. 2002;81(1-3):174–179.

36. Reisberg D. Cognition : exploring the science of the mind. Seventh edition ed. W.W. Norton & Company; 2019.

37. Burke DM, Shafto MA. Aging and language production. Current directions in psychological science. 2004;13(1):21–24.

38. Kang B, Ma J, Jeong I, et al. Behavioral marker-based predictive modeling of functional status for older adults with subjective cognitive decline and mild cognitive impairment: Study protocol. Digital Health. 2024;10:20552076241269555.

39. Kandel ER. Principles of neural science. 5th ed. McGraw-Hill; 2013.

40. Brown MJ, Patterson R. Subjective Cognitive Decline Among Sexual and Gender Minorities: Results from a U.S. Population-Based Sample. J Alzheimers Dis. 2020;73(2):477–487. doi:10.3233/jad-190869

41. Holmen J, Langballe EM, Midthjell K, et al. Gender differences in subjective memory impairment in a general population: the HUNT study, Norway. BMC psychology. 2013;1(1):19.

42. Marinou S, Taler V. Age and sex differences in cognitive performance in people with subjective cognitive decline and associated worry: Findings from the Canadian Longitudinal Study on Aging. Journal of Neuropsychology. 2026;

43. Heser K, Kleineidam L, Wiese B, et al. Subjective cognitive decline may be a stronger predictor of incident dementia in women than in men. Journal of Alzheimer’s Disease. 2019;68(4):1469–1478.

44. Vannini P, Amariglio R, Hanseeuw B, et al. Memory self-awareness in the preclinical and prodromal stages of Alzheimer’s disease. Neuropsychologia. 2017;99:343–349.

45. Joubert S, Gardy L, Didic M, Rouleau I, Barbeau EJ. A meta-analysis of semantic memory in mild cognitive impairment. Neuropsychology Review. 2021;31(2):221–232.

46. Zullo L, Clark C, Gholam M, et al. Factors associated with subjective cognitive decline in dementia□free older adults—A population□based study. International Journal of Geriatric Psychiatry. 2021;36(8):1188–1196.

