## Supplementary materials for "Subjective word-finding complaints in older adults: Development and preliminary psychometric evaluation of a novel self-reported questionnaire"

**Supplementary Figure 1**: Item response distribution of the SWFC questionnaire. The frequency and percentage of participant item endorsement is indicated in each bar

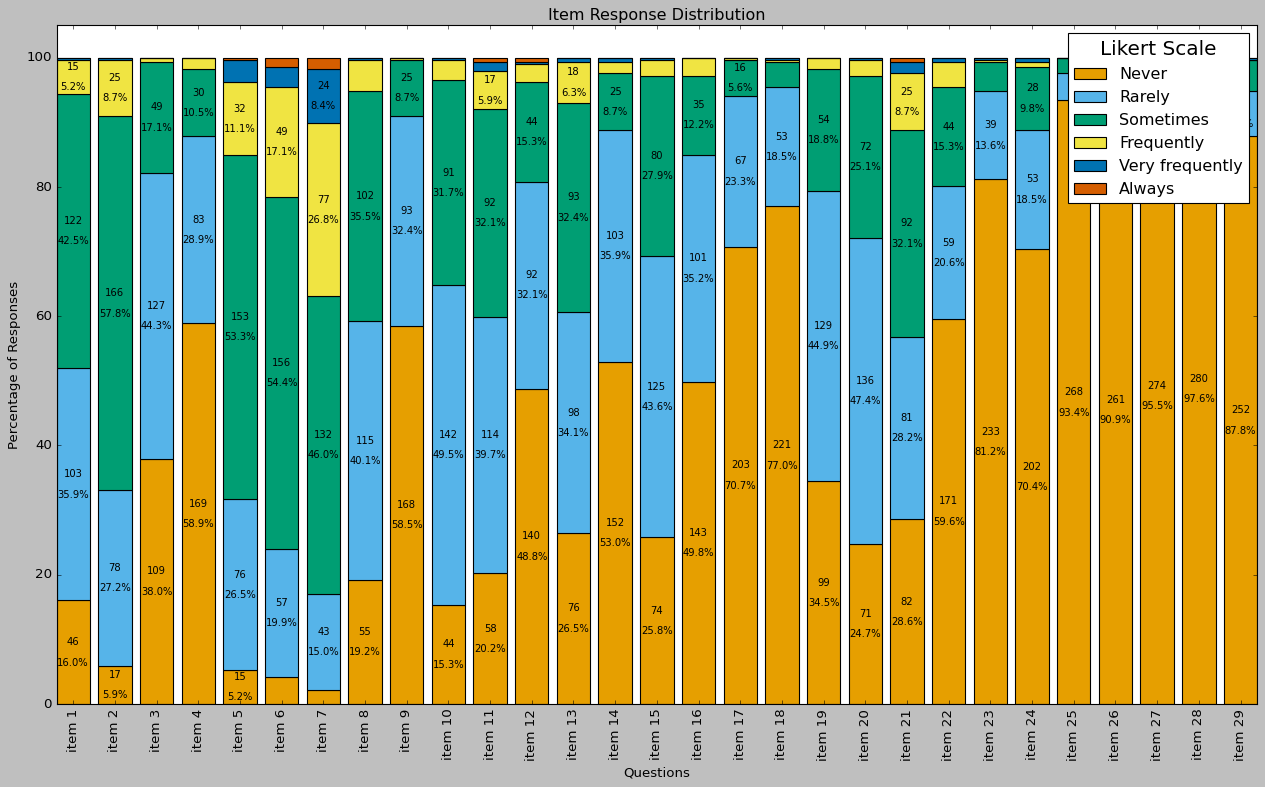

**Supplementary Figure 2**: The inter-item correlation matrix shows Pearson correlation coefficients for all the SWFC questionnaire items. Darker blue shading meant that the items did not correlate well with each other, and darker red shading meant that the items were strongly correlated with each other.

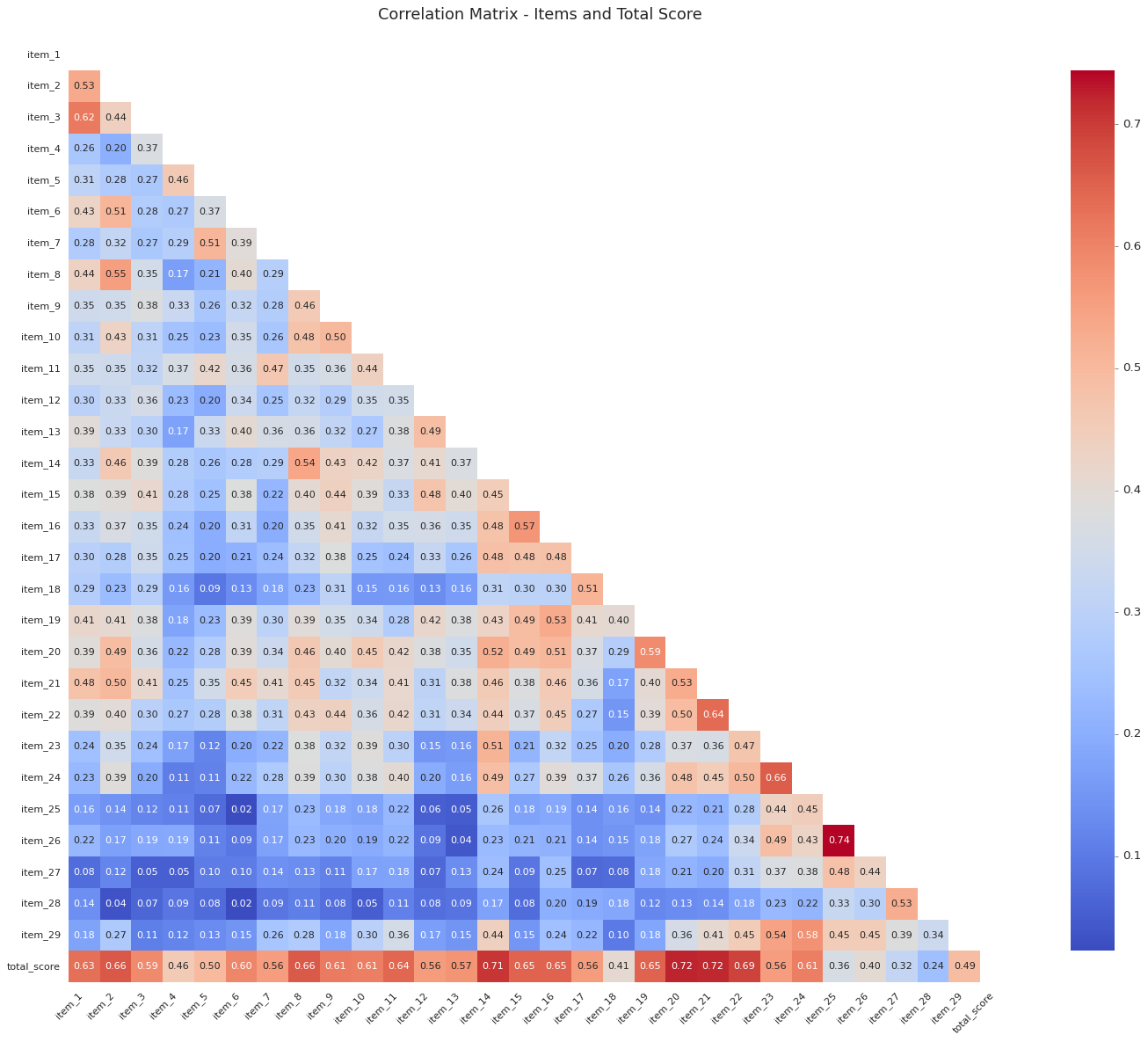

**Supplementary Figure 3**: The normality distribution of the questionnaire (n=29). The bars represent the frequency counts of each total score. The dashed line represents the normal probability density function, plotted for comparison with the observed data. Visual inspection suggests that the distribution is approximately normal with a slight right skew.

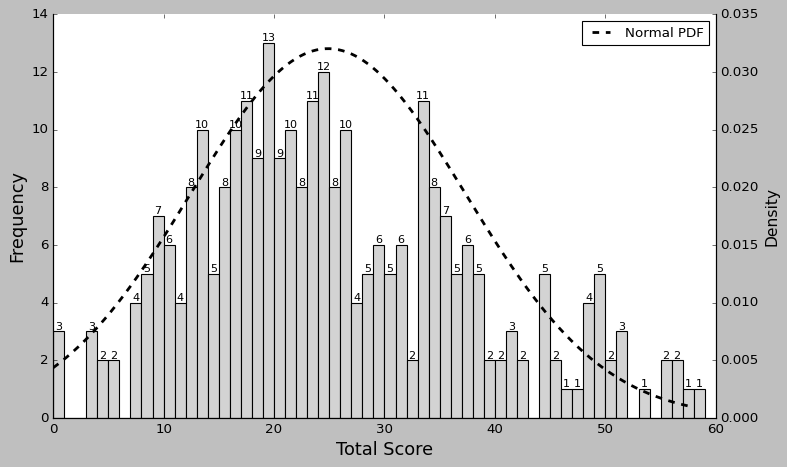

**Supplementary Table 1:** SWFC questionnaire items in French

| **PAR RAPPORT À IL Y A 10 ANS, À QUELLE FRÉQUENCE…** | | 0 | 1 | 2 | 3 | 4 | 5 |
| --- | --- | --- | --- | --- | --- | --- | --- |
|  |  | Jamais | Rarement | Parfois | Fréquement | Très fréquement | Toujours |
| 1 | oubliez-vous les noms d’objets ? |  |  |  |  |  |  |
| 2 | avez-vous de la difficulté à trouver le mot exact à utiliser lors d'une conversation ? |  |  |  |  |  |  |
| 3 | avez-vous du mal à trouver les noms exacts des objets du quotidien couramment utilisés ? |  |  |  |  |  |  |
| 4 | oubliez-vous les noms de vos amis et de votre famille ? |  |  |  |  |  |  |
| 5 | oubliez-vous les noms des personnes que vous connaissez, mais avec lesquelles vous n'êtes pas proches ? |  |  |  |  |  |  |
| 6 | sentez-vous que les mots sont sur le bout de votre langue? |  |  |  |  |  |  |
| 7 | avez-vous de la difficulté à vous rappeler des noms de personnes célèbres (acteurs, politiciens, athlètes, chanteurs, etc.) ? |  |  |  |  |  |  |
| 8 | faites-vous des pauses lors des conversations pour vous rappeler de certains mots ? |  |  |  |  |  |  |
| 9 | est-il difficile pour vous d'écrire des messages textes ou des courriels en raison de difficultés à trouver les mots ? |  |  |  |  |  |  |
| 10 | hésitez-vous avant de choisir le mot exact que vous voulez dire ? |  |  |  |  |  |  |
| 11 | avez-vous du mal à trouver les noms d’endroits ou de villes ? |  |  |  |  |  |  |
| 12 | utilisez-vous votre corps pour vous aider à vous exprimer lorsque vous avez de la difficulté à trouver un mot ? (par exemple, expressions faciales, mouvements des mains) |  |  |  |  |  |  |
| 13 | demandez-vous à votre interlocuteur de vous aider à trouver un mot que vous n’arrivez pas à trouver ? |  |  |  |  |  |  |
| 14 | avez-vous tendance à ne pas terminer vos phrases en raison de votre difficulté à trouver les mots ? |  |  |  |  |  |  |
| 15 | utilisez-vous des mots vagues comme (« quelque chose », « ça », « truc ») au lieu de mots plus précis ? |  |  |  |  |  |  |
| 16 | remplacez-vous un mot spécifique par un mot plus général ? (par exemple, dire « animal » au lieu de « vache ») |  |  |  |  |  |  |
| 17 | remplacez-vous un mot spécifique par un autre mot qui lui est lié ? (par exemple, dire « pomme » au lieu de « poire ») |  |  |  |  |  |  |
| 18 | mélangez-vous les sons dans un mot ? (par exemple, dire « table » au lieu de « sable ») |  |  |  |  |  |  |
| 19 | décrivez-vous un concept au lieu d'utiliser le mot précis ? |  |  |  |  |  |  |
| 20 | avez-vous tendance à tourner autour du pot avant de trouver un mot précis ? |  |  |  |  |  |  |
| 21 | êtes-vous frustré(e) à cause de votre difficulté à trouver les mots ? |  |  |  |  |  |  |
| 22 | êtes-vous triste à cause de votre difficulté à trouver les mots ? |  |  |  |  |  |  |
| 23 | limitez-vous vos conversations à cause de votre difficulté à trouver les mots ? |  |  |  |  |  |  |
| 24 | êtes-vous gêné lors des conversations ou des prises de parole en public en raison de votre difficulté à trouver les mots ? |  |  |  |  |  |  |
| 25 | évitez-vous les activités sociales à cause de votre difficulté à trouver les mots ? |  |  |  |  |  |  |
| 26 | avez-vous du mal à apprécier les activités sociales en raison de votre difficulté à trouver les mots ? |  |  |  |  |  |  |
| 27 | avez-vous du mal à apprécier les activités sociales en raison de votre difficulté à trouver les mots ? |  |  |  |  |  |  |
| 28 | les gens parlent-ils avec vous d'une façon qui semble infantilisante, en sursimplifiant ou en ralentissant excessivement la conversation, à cause de vos difficultés à trouver les mots ? |  |  |  |  |  |  |
| 29 | ressentez-vous de l'anxiété dans les contextes sociaux à cause de votre difficulté à trouver les mots ? |  |  |  |  |  |  |

**Supplementary table 2: Sample demographics stratified by test language**

|  | **French** | **English** | **p-value** | **Whole Sample** |
| --- | --- | --- | --- | --- |
| **Frequency** | 259 | 28 | - | 287 |
| **Sex** (M/F) | 86/173 | 3/25 | 0.026* | 89/198 |
| **Age** (Mean ± SD (Range)) | 71.23 ± 5.52  (56-87) | 74.00 ± 6.27  (57-90) | 0.013* | 71.50 ± 5.64  (56-90) |
| **Years of education^a^** (Mean ± SD (Range)) | 15.78 ± 3.21  (7-26) | 16.48 ± 4.72  (9-29) | 0.722 | 15.85 ± 3.38  (7-29) |
| **Cognitive status** (CU/MCI) | 174/85 | 18/10 | 0.922 | 192/95 |
| **SWFC Total Score** (Mean ± SD (Range)) | 24.50 ± 12.37  (0-57) | 28.50 ± 12.94  (5-58) | 0.104 | 24.89 ± 12.46  (0-58) |
| **MMSE scores^b^** (mean ± SD (Range)) | 28.74 ± 1.37  (22-30) | 28.64 ± 1.89  (22-30) | 0.762 | 28.73 ± 1.43  (22-30) |
| **RBANS total score^c^** ((mean ± SD (Range)) | 99.71 ± 10.02 (70-125) | 101.76 ± 11.47 (78-127) | 0.476 | 99.93 ± 10.17 (70-127) |
| **RBANS Delayed Memory Recall^d^** ((mean ± SD (Range)) | 102.41 ± 9.54 (56-127) | 105.52 ± 9.30 (81-122) | 0.137 | 102.75 ± 9.54 (56-127) |
| **RBANS language index score^e^** (mean ± SD (Range)) | 98.98 ± 9.62 (68-134) | 101.86 ± 10.75 (83-124) | 0.195 | 99.29 ± 9.76 (68-134) |
| **Multilingualism^f^** (monolingual/multilingual) | 50/142 | 3/20 | 0.267 | 53/162 |

*Notes:* a. Years of education was available for n=285; b. MMSE scores were available for n=198. c. RBANS total scores were available for n=195; d. RBANS delayed memory recall scores were available for n=197; e. RBANS language index scores were available for n=197; f. Multilingualism was available for n=215.

**Supplementary Table 3:** Effects of demographics and clinical factors on SWFC factor scores.

| **SWFC Factor** | **Predictor** | **F** | **df** | **p-value** | **Partial effect size (η²)** |
| --- | --- | --- | --- | --- | --- |
| **Factor 1. SWFC coping strategies** | Age | 3.80 | 1, 279 | 0.052 | 0.013 |
|  | Sex | 1.78 | 1, 279 | 0.183 | 0.006 |
|  | Education | 0.03 | 1, 279 | 0.863 | 0.000 |
|  | Cognitive status | 0.02 | 1, 279 | 0.884 | 0.000 |
|  | Multilingualism | 0.07 | 1, 177 | 0.799 | 0.000 |
| **Factor 2. Fluency difficulties and affective impacts** | Age | 2.95 | 1, 279 | 0.087 | 0.010 |
|  | Sex | 0.28 | 1, 279 | 0.594 | 0.001 |
|  | Education | 1.13 | 1, 279 | 0.289 | 0.004 |
|  | Cognitive status | 1.87 | 1, 279 | 0.173 | 0.007 |
|  | Multilingualism | 0.40 | 1, 177 | 0.527 | 0.002 |
| **Factor 3. SWFC-related errors** | Age | 3.20 | 1, 279 | 0.075 | 0.011 |
|  | Sex | 1.56 | 1, 279 | 0.212 | 0.006 |
|  | Education | 2.37 | 1, 279 | 0.125 | 0.008 |
|  | Cognitive status | 0.00 | 1, 279 | 0.948 | 0.000 |
|  | Multilingualism | 1.06 | 1, 177 | 0.306 | 0.006 |
| **Factor 4. Proper noun retrieval difficulties** | Age | 6.90 | 1, 279 | 0.009** | 0.024 |
|  | Sex | 7.28 | 1, 279 | 0.007** | 0.025 |
|  | Education | 0.05 | 1, 279 | 0.829 | 0.000 |
|  | Cognitive status | 0.79 | 1, 279 | 0.374 | 0.003 |
|  | Multilingualism | 0.86 | 1, 177 | 0.355 | 0.005 |
| **Factor 5. Social impacts of SWFC** | Age | 2.89 | 1, 279 | 0.090 | 0.010 |
|  | Sex | 0.04 | 1, 279 | 0.848 | 0.000 |
|  | Education | 4.21 | 1, 279 | 0.041* | 0.015 |
|  | Cognitive status | 6.28 | 1, 279 | 0.013* | 0.022 |
|  | Multilingualism | 0.38 | 1, 177 | 0.537 | 0.002 |
| **Factor 6. Common noun retrieval difficulties** | Age | 7.80 | 1, 279 | 0.006** | 0.027 |
|  | Sex | 1.54 | 1, 279 | 0.215 | 0.005 |
|  | Education | 0.09 | 1, 279 | 0.763 | 0.000 |
|  | Cognitive status | 0.30 | 1, 279 | 0.582 | 0.001 |
|  | Multilingualism | 0.01 | 1, 177 | 0.939 | 0.000 |
